# Improving population-scale disease prediction through multi-omics integration

**DOI:** 10.1101/2025.11.24.25340920

**Authors:** Esther S. Ng, Ruth Nanjala, Luke Jostins-Dean, Jeffrey C. Barrett, John A. Todd, Yang Luo

## Abstract

The early detection of common diseases is currently constrained by a dependence on single biomarkers, which often capture only a limited aspect of disease pathology. Here, we applied multi-omic factor analysis to integrate plasma proteins, metabolites, biochemical and haematological measurements from 46,081 individuals in the UK Biobank Pharma Proteomics Project. We identified 20 latent factors that capture major axes of biological variation across omic layers and found that these factors showed 400 significant associations with 111 incident diseases. We then incorporated the top 50 multi-omic features per factor into predictive models and observed improved disease discrimination for 85% of conditions compared with the best single biomarker. For example, the multi-omic models for predicting incident diabetes (C-index = 0.865, 95%CI: 0.850-0.882) and iron-deficiency anaemia (C-index = 0.750, 95%CI: 0.726 - 0.778) significantly outperformed their respective clinical biomarkers, HbA1c (C-index = 0.810, 95%CI: 0.791-0.831) and haemoglobin concentration (C-index = 0.668, 95%CI: 0.644-0.692). We therefore conclude that population-scale multi-omic integration reveals shared disease mechanisms and enhances risk prediction, providing a framework for earlier and biologically informed diagnosis and prevention.

## Main

Current approaches to disease diagnosis rely predominantly on laboratory tests that detect cellular or molecular abnormalities. While these clinical assays are essential, they often provide only a partial view of the underlying biology. Most complex diseases involve disruptions across multiple biological systems, including metabolic, immune and genetic factors that are not fully captured by single-feature tests. As a result, opportunities for early diagnosis, risk stratification and individualized treatment are frequently missed.

Multi-omic data integration, that is, the combined analysis of diverse molecular layers such as proteomics, metabolomics, transcriptomics and genomics, has emerged as a promising strategy to address this limitation. By capturing a more complete view of biological states, multi-omics enables the identification of latent disease signatures and novel mechanistic insights^1–3^. However, large-scale, population-level applications of multi-omic profiling remain relatively limited.

The recent advent of high throughput metabolomic and proteomic technology has enabled large-scale profiling of metabolites and proteins at the population level. In this study, we used deep molecular phenotyping data from 46,081 participants in the UK Biobank Pharma Proteomics Project (UKB-PPP), one of the largest prospective health cohorts globally. We integrated proteomic^4^, metabolomic^5^, clinical biochemistry, haematology and electronic health record data to generate a comprehensive multi-omic landscape of common diseases. Our objectives are: (1) to systematically map multi-omic features associated with a wide range of common diseases; (2) to identify genetic variants that regulate these disease-associated multi-omic modules; and (3) to evaluate whether multi-omic feature sets can improve diagnostic performance compared to individual clinical assays.

## Results

### Feature selection and multi-omic integration

We hypothesized that integrating multiple data types could reveal novel multicellular features in complex diseases and link these features to disease outcomes. To test this, we applied Multi-Omic Factor Analysis (MOFA)^2^ to data from the UKB-PPP, which includes proteins, metabolites, biochemical markers and haematological measures from 46,081 participants (**Fig.1**). MOFA provides an unsupervised framework for exploring relationships across diverse molecular data types by inferring a low-dimensional representation in the form of latent factors. These latent factors capture the primary axes of variation across data modalities, facilitating the identification of shared underlying molecular processes and disease subgroups.

**Fig. 1:**
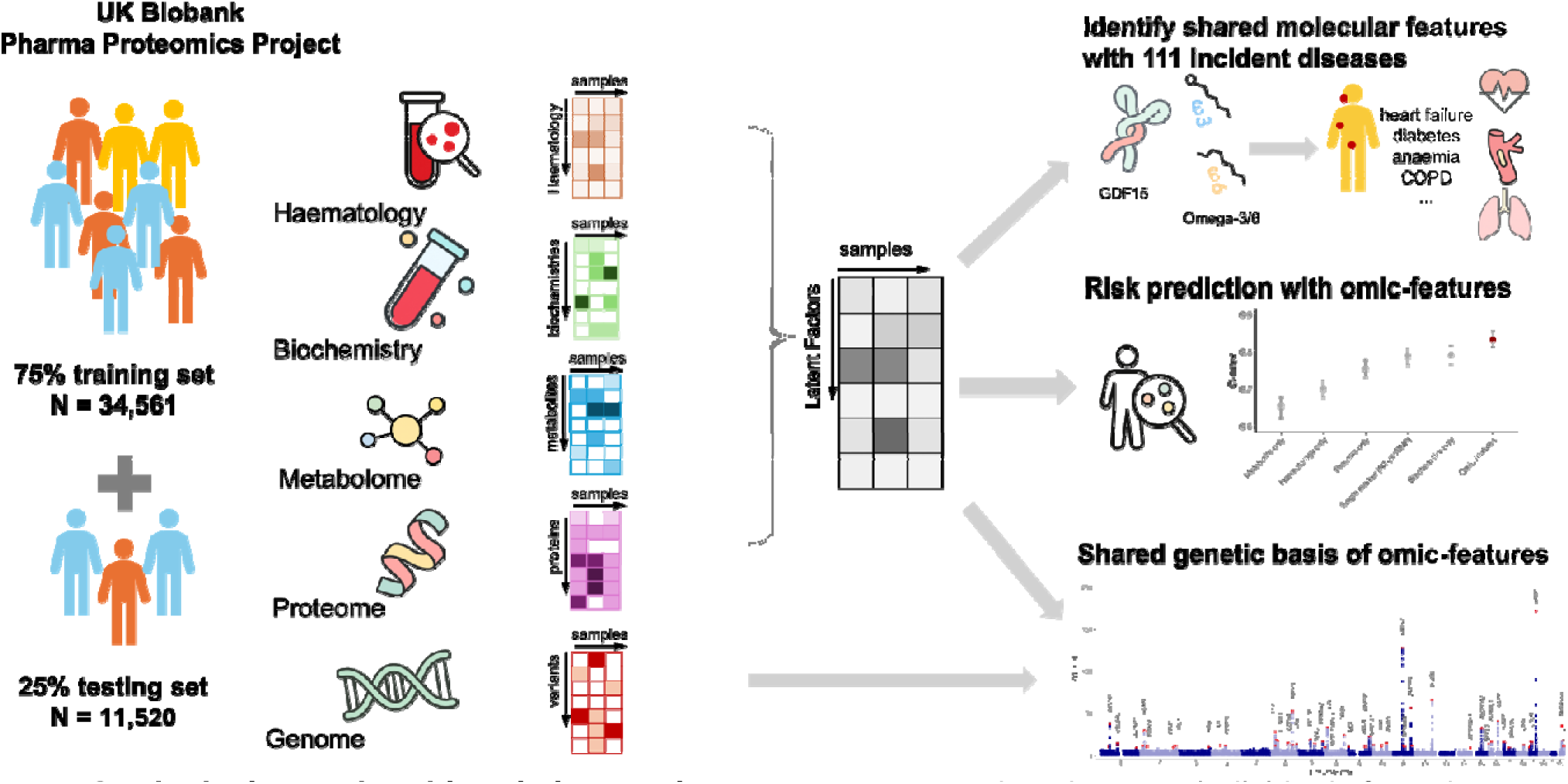
Study design and multi-omic integration strategy. We analyzed 46,081 individuals from the UK Biobank with matched proteomic, metabolomic, biochemical and haematological measurements. Participants were randomly split into a training set (N = 34,561; 75%) for feature selection and model development, and a testing set (N = 11,520; 25%) for validation and replication. After feature selection across modalities, a Multi-Omic Factor Analysis (MOFA) model was trained in the training cohort to infer latent factors representing major axes of molecular variation. Downstream analyses included latent factor characterization, factor-disease association, genetic association analysis (factor-GWAS) and disease prediction. All MOFA-derived features and predictive models were evaluated in the he d-out testing set to ensure robustness and generalizability.

We began by curating a subset of features from each data type to balance biological relevance and computational tractability. There is a total of 249 metabolites measured via nuclear magnetic resonance spectroscopy in blood samples across all UKB individuals^5^ (N = 477,706). We observed an extensive correlation structure among these metabolites (**Supplementary Fig.1**). We selected 36 key metabolites for MOFA analysis based on their clinical utility and ability to predict disease^5^.

We included 19 and 22 features for biochemical markers and haematology values, respectively, after removing duplicated or derivative values already captured by the metabolomics or proteomics platforms. Specifically, we excluded percentage-based haematology metrics, retaining only absolute counts for consistency and interpretability.

To select a manageable yet informative subset of proteins, we performed supervised feature selection from the 2,920 unique proteins measured in the UKB-PPP^4^. Using curated and validated phenotypes for 111 incident diseases^6^, we trained LASSO regression models for each disease, adjusting for age and sex, in a randomly selected 75% subset of participants with proteomic data (N = 34,561). We included only incident diseases occurring after biospecimen collection and with at least 100 cases in the training dataset (**Supplementary Table 1**). For each disease, we selected the top three proteins most strongly associated with disease risk, ranked by absolute effect size (/3), resulting in a panel of 217 unique proteins for MOFA integration.

In total, we integrated 217 proteins, 36 metabolites, 19 biochemical markers and 22 haematology features from 34,561 individuals for MOFA training (**Supplementary Table 2**), holding out the remaining 25% of individuals (N = 11,520) for downstream validation (**Fig. 1**).

We identified 20 latent factors with each factor explaining a minimum of 1.5% variance. As expected, we saw that most of these factors captured variation shared across multiple data types (**Fig. 2a**), supporting the presence of coordinated biological processes spanning molecular domains. Cumulatively, these 20 factors explained 81.8% of the variance in metabolites, 30.8% in biochemistry, 36.8% in proteomics and 35.2% in haematology (**Fig. 2b**). When examining associations with demographic and anthropometric characteristics, we found that several factors were strongly correlated with age, sex and BMI (**Supplementary Fig. 2**). For instance, top weighted features in Factor 2 included established clinical biomarkers such as HDL^7^, sex hormone binding globulin (SHBG)^8^ and haemoglobin concentration^9^ (**Fig. 2c**). Consequently, it is associated with sex, BMI and BMI-related disease outcomes including diabetes (**Fig. 2d-f**). These findings validate the biological relevance of our multi-omic factor structure and demonstrate the potential of unsupervised integration to uncover clinically meaningful axes of variation at population scale.

**Fig. 2.**
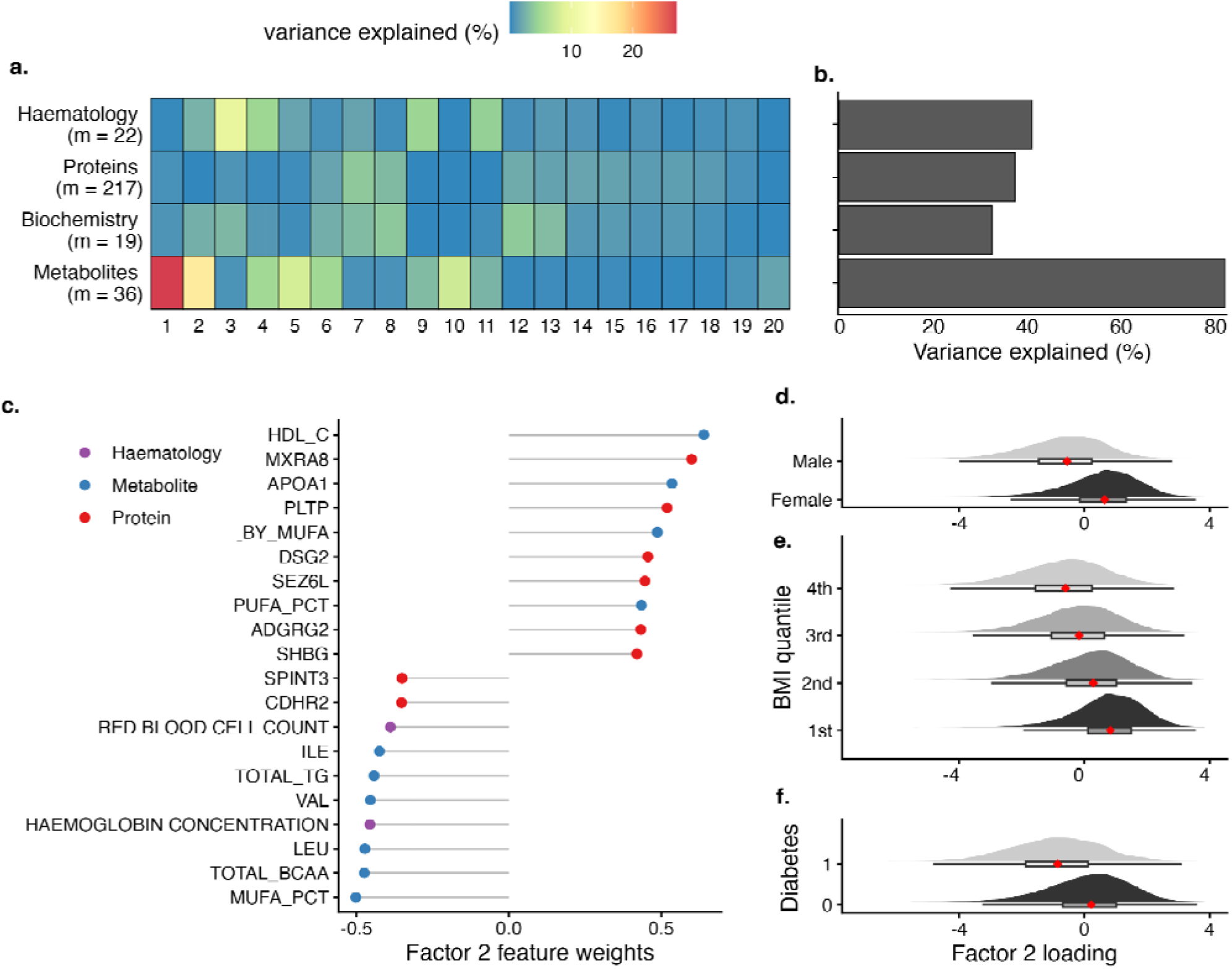
Overview of multi-omic integration and latent factor characterization. **a**. Variance decomposition showing the proportion of total variance explained (R^2^) by each of the 20 latent factors across individual data modalities. **b**. Cumulative proportion of total variance explained by all 20 factors in each data modality. **c**. Top 10 positive and top 10 negative feature loadings of Factor 2. Distribution of Factor 2 scores **d**. stratified by sex (female vs. male). **e.** across quartiles of BMI. **f**. in individuals with or without diabetes. In (**e–f**), boxplots represent the 25th and 75th percentiles, with whiskers extending from minimum to maximum values. Red dots indicate the median Factor 2 value. Density plots next to each boxplot show the distribution of Factor 2 loadings per category.

### Latent factors have strong pleiotropic disease associations

To evaluate the clinical relevance of the latent factors inferred from multi-omic integration, we assessed their associations with disease risk. Specifically, we performed logistic regression analyses of each of the 20 latent factors against 111 incident diseases, adjusting for age and sex. We did not adjust for BMI because there are various metabolic diseases which are mediated by BMI and several of the metabolites (such as cholesterol and triglycerides) are clearly associated with BMI. Therefore, we reasoned adjusting for BMI would attenuate or extinguish disease associations that are driven by BMI. In total, we found 400 unique incident disease-factor associations after a Bonferroni-corrected significant threshold corrected for multiple testing (0.05/(111×20), **Fig. 3a**, **Supplementary Table 3**). Consistent with our feature selection strategy for prioritizing disease-relevant molecular features, we noted that all latent factors were associated with multiple diseases, revealing broad molecular pleiotropy spanning distinct clinical categories (**Supplementary Fig. 3a**). Notably, for most latent factors, disease associations showed a consistent direction of effect across traits (**Fig. 3b, Supplementary Fig. 4**).

**Fig. 3.**
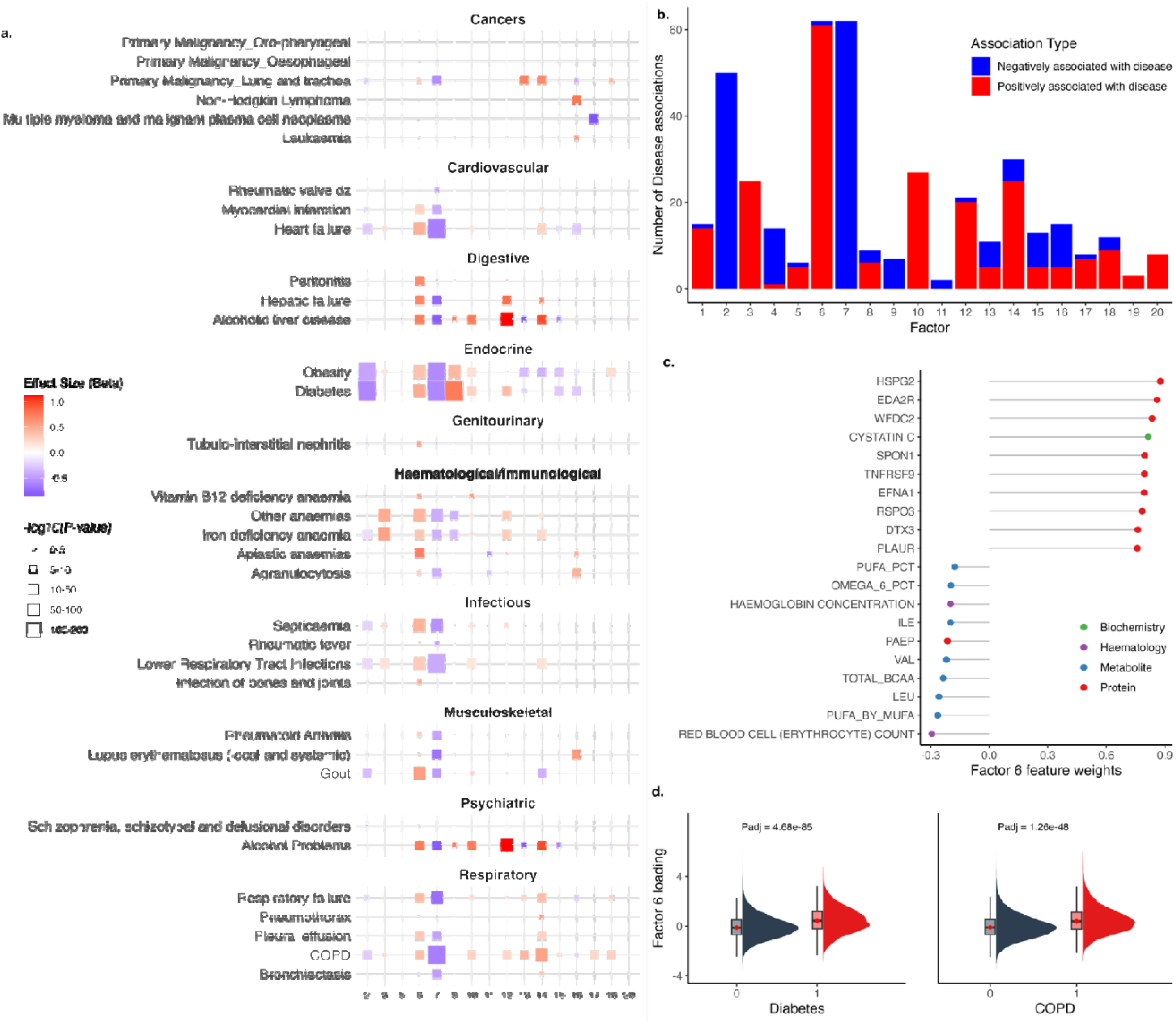
Latent factors show widespread and pleiotropic associations with disease. **a**. Factor-disease associations for 111 incident diseases in the UKB-PPP, estimated via logistic regression adjusted for age and sex. For visualization, only factors with a Bonferroni-corrected P-value < 0.05 and an odds ratio > 1.5 or < 2/3 for any disease are shown. Tiles are colored based on association strength (regression), with size proportional to -log10(P-value). Full results are shown in **Supplementary** Fig. 4. and included in **Supplementary Table 3**. **b**. Histogram showing the number of disease associations per factor, considering only disease-factor associations with a Bonferroni-corrected P-value < 0.05. Bars are colored by whether the factor is positively (red) or negatively (blue) associated with disease prevalence. **c**. Factor loadings of top 10 positive and top 10 negative features in Factor 6. **d**. Distribution of Factor 6 loadings in individuals with and without diabetes (= 0.491, P_adj_ = 4.68×10^-85^) and COPD (= 0.390, P_adj_ = 1.26×10^-48^). Boxplots represent the 25^th^ and 75^th^ percentiles, with whiskers extending from minimum to maximum values. Red dots indicate the median value of Factor 6 loadings.

Among these, Factors 6 and 7 stood out as the most disease-associated (62/111), yet they represent distinct molecular axes (**Supplementary Fig. 3b-c**). looking into the features with the highest loadings in Factor 6 (**Fig. 3c**), it integrates hematological, metabolic and protein features characteristic of a metabolic–inflammatory axis. Top loadings such as haemoglobin concentration and branched-chain amino acids (leucine, isoleucine, valine), mark altered oxygen transport and amino-acid metabolism linked to insulin resistance and type 2 diabetes^10^. Lipid traits such as ratio of polyunsaturated fatty acids (PUFA) to monounsaturated fatty acids (MUFA), omega-6 percentage, total PUFA percentage, indicate disrupted fatty-acid balance associated with obesity, cardiometabolic and respiratory disease^11^. Consistent with this profile, Factor 6 was positively associated with diabetes (/3 (se) = 0.491 (0.0246), P_adj_ = 4.68 × 10^-85^, **Fig. 3d**), obesity (/3 (se) = 0.359 (0.0204), P_adj_ = 7.66 × 10^-66^), heart failure (/3 (se) = 0.457 (0.0272), P_adj_ = 5.03 × 10^-60^) and COPD (/3 (se) = 0.390 (0.0257), P_adj_ = 1.26 × 10^-48^, **Fig. 3e**). In contrast, Factor 7 is predominantly protein-based and exhibits inverse associations with many diseases. Its leading proteins including LGALS9, ADM, VSIG4, LEP, CRP and GDF15 represent immune-regulatory and stress-response pathways that promote vascular and metabolic homeostasis^12–15^. Notably, Factor 7 was inversely associated with obesity (/3 (se) = −0.570 (0.0213), P_adj_ = 1.44 × 10^-154^) and diabetes (/3 (se) = −0.586 (0.0257), P_adj_ = 3.37 × 10^-122^), suggesting a protective, protein-mediated axis opposing the risk-enriched metabolic signature of Factor 6.

Diseases with the greatest number of factor associations (16/20) are diabetes and obesity (**Supplementary Fig. 5**). This is consistent with the known widespread impact of both conditions on metabolic and proteomic profiles^16–18^.

To validate the robustness of these associations, we projected the trained MOFA model into the held-out 25% of participants (N = 11,520) and repeated the logistic regression analyses. Factor values were reconstructed in the validation set using the trained MOFA loadings and associations with disease outcomes were reassessed. We observed a high degree of replication, with 98.5% (394 of 400) of the discovery associations showing a concordant direction of effect in the validation cohort (**Supplementary Fig. 6**).

### Genome-wide association studies of latent factors

We next examined the genetic basis of the multi-omic latent factors by performing genome-wide association studies (GWAS) on the MOFA-inferred factor weights. We hypothesized that these latent traits capture biologically relevant, genetically regulated axes of molecular variation that may underlie complex diseases.

To this end, we conducted GWAS for each of the 20 latent factors within the MOFA training set, restricting our analysis to individuals of white British ancestry (N□=□28,693; UKB Field ID: 22006), adjusting for age, sex and the first 10 genetic principal components in the regression models (**Fig. 4a**). This factor-GWAS approach allowed us to investigate the common variant architecture contributing to each inferred latent factor. We examined genomic inflation factors (*λ_gc_*) of each of the 20 factor-GWAS and observed no inflation of test statistics (median *λ_gc_* = 1.05, **Supplementary Fig. 7**), suggesting that our model is robust to false-positive findings.

**Fig. 4.**
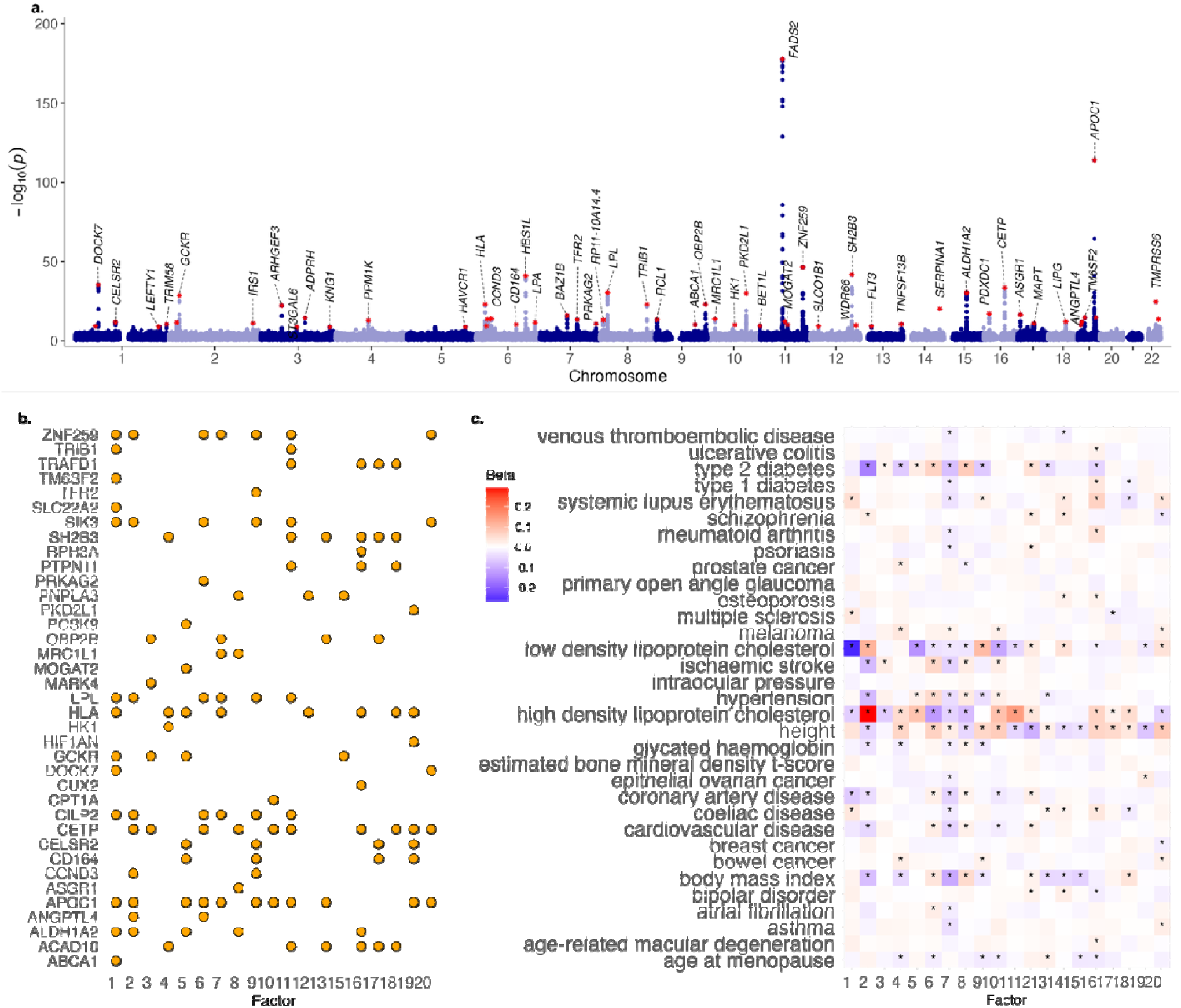
Factor-GWAS of MOFA-derived latent factors. **a**. Manhattan plot summarizing genome-wide association results for each of the 20 MOFA-inferred latent factors. The x-axis indicates genomic location, whereas the y-axis shows the (-log_10_(P)) from the linear regression model. The red dots how lead SNP for each independent locus passing the Bonferroni-correct threshold (5×10^-8^/20) and are labelled with the name of the nearest protein-coding gene. **b.** The number of factors associated with the 37 factor-associated genes. **c**. Heatmap showing associations between polygenic risk scores (PRS) for 36 complex traits and the weights of 20 latent factors. Each cell represents the effect size () from linear regression adjusted for age and sex. Asterisks denote Bonferroni-significant associations (P□<□0.05/720).

We identified 37 independent loci significantly associated with at least one latent factor (**Supplementary Table 4**). We found that 22/37 loci were associated with at least two latent factors, including known pleiotropic regions such as the *HLA* and *APOC1* locus (**Fig. 4b**), suggesting shared genetic regulation across multiple biological processes.

To investigate the biological pathways underpinning these genetic associations, we mapped per-factor GWAS summary statistics to gene-level associations and performed gene set enrichment analysis using Multi-marker Analysis of GenoMic Annotation^19^ (MAGMA) with REACTOME pathway annotations^20^. We found that many factor-loci were enriched in lipid metabolism pathways, including chylomicron remodeling, clearance and assembly, which were associated with multiple factors (**Supplementary Fig. 8**). These findings align with previously identified factor-disease associations, suggesting that many of these factors are linked to metabolite-associated diseases, further supporting their biological relevance.

We further assessed the relevance of the factor-GWAS loci by comparing them to the NHGRI-EBI GWAS Catalog^21^. Of the 37 independent loci, 36 (97.3%) had been previously reported in association with at least one phenotype (**Supplementary Table. 5**). Many of these SNPs were highly pleiotropic suggesting that there may be common pathways mediated through these factors. An example is rs1260326, which is a missense variant in *GCKR* gene and significantly associated with Factors 1, 3, 5 and15 (Bonferroni-corrected P-value < 0.05). This variant has previously been implicated in diverse traits including liver enzymes, C-reactive protein, urate levels, lipids, calcium, creatinine, platelet count, sex hormone-binding globulin, IGF-1 and diseases such as gout, Crohn’s disease, psoriasis and ankylosing spondylitis^22–25^

To ensure the robustness of our factor-GWAS results, we projected the trained MOFA weights onto the 25% held-out test set and re-performed the factor-GWAS analyses. All 37 lead SNPs identified in the 75% discovery set exhibited directionally concordant effects in the test set (**Supplementary Fig. 9**, **Supplementary Table 6**), providing strong evidence that the inferred loci are reproducible and not driven by sampling variation.

Finally, to evaluate whether genetically inferred disease risk is reflected in the MOFA-derived factors, we assessed associations between latent factors and polygenic risk scores (PRS) for 36 traits computed by Genomics PLC for UKB participants^26^ (**Supplementary Table 7**). We performed linear regression models between PRS and factor-weights adjusted for age and sex. We observed 174/720 significant PRS–factor associations after Bonferroni correction (P□<□0.05/720; **Fig. 4c**). The strongest association was between Factor 2 and the PRS for HDL cholesterol, consistent with HDL being a top-loading feature in Factor 2 and this factor’s association with metabolic syndrome traits including diabetes, obesity, and hypertension.

Multiple other factors were associated with PRS for LDL cholesterol, BMI, type 2 diabetes, cardiovascular disease, and coronary artery disease, further reinforcing the clinical relevance of these multi-omic dimensions.

### Omic-features improve disease prediction over single modality

To evaluate the predictive utility of multi-omic latent factors, we assessed their ability to discriminate between individuals with and without disease using the concordance index (C-index) across 80 of 111 incident diseases that showed significant associations with at least one MOFA factor. The C-index ranges from 0 (no discrimination) to 1 (perfect discrimination). For each factor, we ranked features by their absolute loadings and retained the top 50 features to train logistic regression models adjusted for age and sex. Models were fitted in a 75% training cohort and evaluated in the remaining 25% using 100 bootstrap replicates (5,000 samples per replicate). To ensure sufficient power, we restricted the analysis to diseases with ≥400 incident cases, yielding a final set of 53 diseases for prediction (**Supplementary Table 1**).

To benchmark our multi-omic models, we compared their performance against models trained on individual features. For each disease, we evaluated logistic regression models using each of 3,230 single features, including 2,920 proteins, 249 metabolites, 30 biochemical and 31 haematological measures, regardless of whether they were included in MOFA training. This allowed us to assess whether integrating an omic-model yields greater predictive power than any single biomarker.

We found that for 48 of 53 diseases the 50-feature omic-model achieved a higher mean C-index than the best-performing single-feature model (90.6%, **Supplementary Fig. 10a**, **Supplementary Table 8**). Among these, we found six diseases in the lower bound of the 95% confidence interval (CI) for the omic-model that exceeded the upper bound of the best single-feature model, indicating statistically significant improvement in predictive accuracy (**Fig.5a,b**).

One example is iron-deficiency anaemia, where our model based on Factor 9 achieved a C-index with a lower 95% confidence bound of 0.715, exceeding the upper 95% CI of the best single feature and clinically used biomarker, haemoglobin concentration (95% CI: 0.644-0.692). While haemoglobin-related traits such as mean corpuscular haemoglobin and red blood cell count were the highest-weighted features within Factor 9 (**Fig. 5e**), the inclusion of signals from other molecular layers, including omega-3 fatty acids and FGF23, improved predictive performance. FGF23 is a bone-derived hormone that regulates phosphate and vitamin D metabolism, increases in response to iron deficiency and anaemia, reflecting a physiological link between mineral and erythropoietic pathways^27,28^. The integration of such cross-system markers, together with lipidomic features that influence red-cell membrane fluidity and deformability^29^, demonstrates how multi-omic models can capture complementary biological signals and enhance disease prediction beyond conventional single biomarkers.

**Fig. 5.**
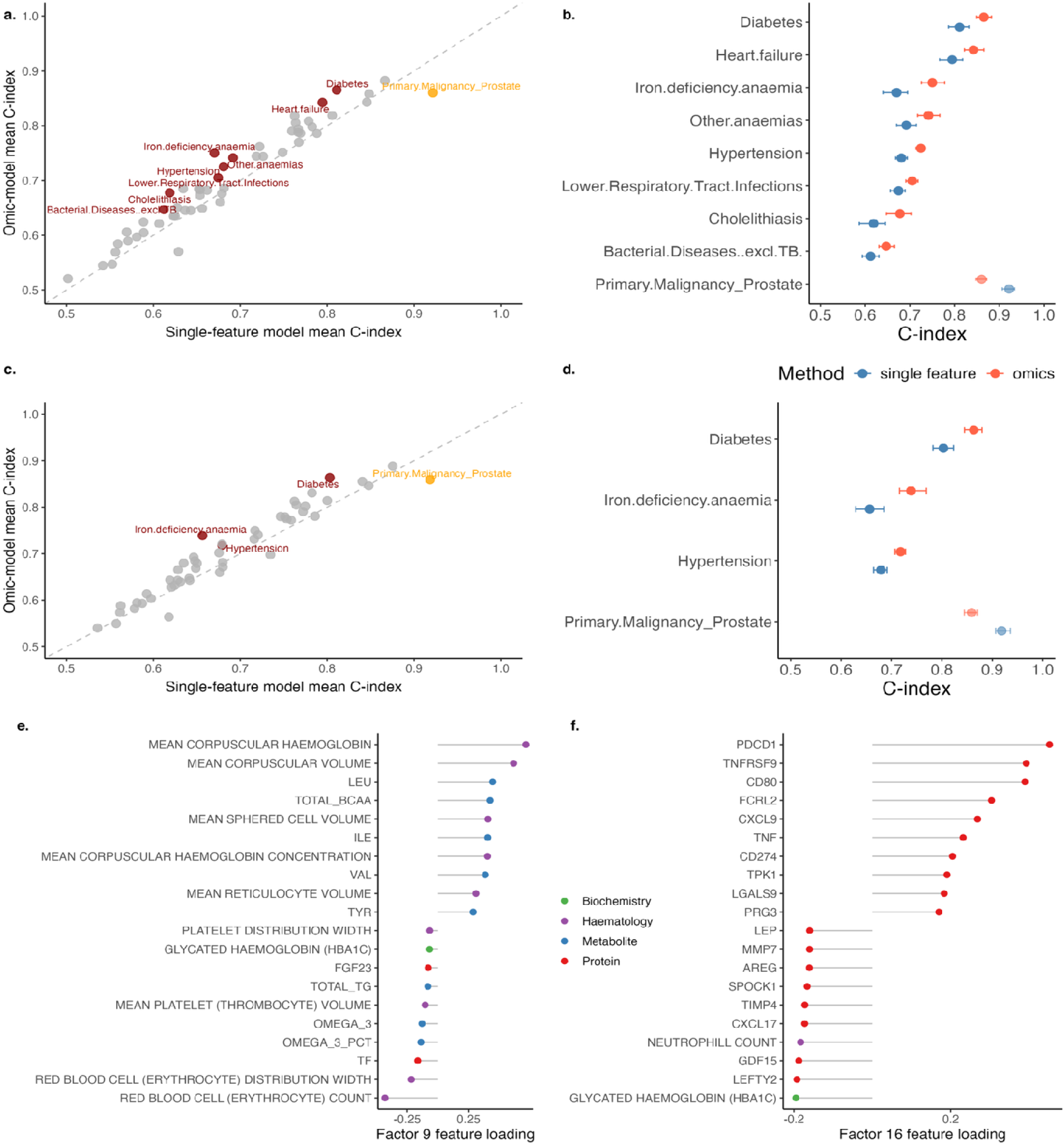
Improved predictive performance of common diseases using multi-omic models compared to single-biomarker models. Scatter plots compare mean C-index for omic-based (y-axis) versus single-biomarker (x-axis) models in (**a**) all-time incident prediction and (**c**) two-year prediction. Each point represents one disease with at least 400 cases. Red points indicate diseases for which the omic-based model significantly outperformed the single-biomarker model, defined as non-overlapping 95% confidence intervals (CIs). The orange point indicates a disease where the single-biomarker model outperformed the omic-based model. Grey points indicate no significant difference. Forest plots showing diseases with non-overlapping 95% confidence intervals between multi-omic (red) and single-feature (blue) models in (**b**) all-time prediction and in (**d**) two-year prediction. Error bars represent 95% confidence intervals around mean C-index values. Top 10 positive and negative feature loadings for factors with superior predictive performance in (**e**) Factor 9 and (**f**) Factor 16. Points are colored by omics modality: metabolites (blue), biochemistry (green), proteins (red), and haematology (purple). These factors demonstrate the multi-omic signatures underlying improved disease prediction performance.

To evaluate whether multi-omic models can predict long-term disease risk, we repeated the analyses using only incident disease cases that occurred at least two years after baseline blood sampling (**Supplementary Table 9**). Among 50 diseases with sufficient case numbers (≥400), 42 showed a higher mean C-index than the best performing single-feature model (84.0%, **Supplementary Fig. 10b**, **Supplementary Table 10**) and three showed significantly improved predictive performance with the factor-based model (**Fig. 5c,d**).

For diabetes, Factor 16 consistently outperformed single biomarkers, achieving a 95% CI C-index of 0.845-0.880, compared with 0.782-0.823 for the clinical biomarker, glycated haemoglobin (HbA1c). This factor was enriched for established diabetes biomarkers such as glucose and HbA1c, alongside several regulatory proteins including GDF15, LEFTY2, and LEP (**Fig. 5f**). GDF15, a stress-induced cytokine that modulates appetite, mitochondrial function, and metabolic homeostasis, has been linked to insulin resistance, weight regulation, and cardiovascular complications in diabetes^30^. Its co-loading with immune mediators such as CXCL9, TNFRSF9, and PDCD1 suggests that Factor 16 captures a metabolic–immune axis underlying progressive diabetes risk. These results indicate that integrating multi-layered omic data enhances both near-term and long-term disease prediction, uncovering mechanistic pathways not evident from single-modality biomarkers.

## Discussion

In this study, we present a comprehensive multi-omic integration framework using MOFA to identify latent factors representing major axes of biological variation across proteomic, metabolomic, biochemical and haematological data in a large, well-characterized population cohort. These latent factors capture shared covariance across omic layers and reveal robust associations with a broad spectrum of common diseases, including metabolic, cardiovascular, renal, neurological diseases and cancer. We observed that several factors also associate with common genetic variants and polygenic risk scores (PRSs) for complex diseases. These associations suggest that the latent factors may act as intermediate molecular phenotypes through which genetic variation influences disease risk. The concordance between factor-disease associations and disease-linked PRSs or GWAS loci highlights the potential of this framework to uncover genetically anchored molecular mechanisms underlying common disease traits.

A key finding from our predictive modeling is that multi-feature models derived from factor loadings can outperform models based on individual features from any single omic modality. This was particularly evident in diseases such as heart failure and anaemia, where our models surpassed the clinical gold-standard biomarkers in discriminatory power. Moreover, in a two-year incident disease prediction analysis, several latent factor-based models retained superior performance, suggesting clinical utility in prospective risk stratification. These results highlight the potential for factor-derived multi-omic signatures to enhance early detection and preventive intervention for diseases with modifiable risk profiles, such as diabetes and cardiovascular disorders.

This study has several limitations. First, MOFA captures only linear relationships between features and may fail to capture important non-linear interactions. Alternative methods that model complex dependencies may provide complementary insights. Second, the accuracy of phenotype definitions in large-scale biobanks such as UK Biobank is limited by diagnostic coding and disease onset may precede recorded diagnosis, introducing potential misclassification. Third, while our models adjust for common confounders, residual confounding remains possible, particularly from unmeasured exposures such as medication use, lifestyle or subclinical disease. Fourth, the interpretability of latent factors remains a challenge. Factors frequently associated with multiple traits including age, sex and BMI, complicating causal interpretation. In some cases, the observed omic correlations may reflect downstream consequences of disease or shared environmental exposures rather than primary etiological mechanisms. Finally, the model’s predictive performance, though encouraging, requires external validation in independent cohorts and diverse populations to assess generalizability and clinical utility.

Together, these findings demonstrate that latent factors derived from integrated multi-omic data can uncover biologically meaningful axes of variation associated with genetic risk and disease. MOFA provides a powerful framework for mechanistically informed disease stratification and enhances prediction beyond single-modality biomarkers. Future work should focus on improving interpretability, modelling non-linear relationships and validating these features in prospective clinical settings to enable their translation into routine risk assessment and precision medicine.

## Methods

### Study design and participants

The UK Biobank (UKB) is a population-based prospective cohort of 502,205 participants residing in the United Kingdom, with extensive phenotypic and genotypic data available. For this study, we focused on the subset of participants from the UK Biobank Pharma Proteomics Project (UKB-PPP) with available Olink Explore proteomics data at baseline (N = 46,081).

For metabolomics, we accessed data on 249 circulating metabolites measured by the Nightingale Health NMR platform in 482,015 participants (Data Field ID:220). Metabolite values exceeding 4 standard deviations from the mean were set to missing. Because alanine values vary by spectrometer, we normalized these per device by centering and scaling within each spectrometer group. All metabolite values were subsequently transformed using log1p normalization. For the proteomics data, we obtained 2,923 proteins from the UKB-PPP^4^ (Data Field ID: 30900). We excluded individuals with more than 50% missing protein values (N = 6,174) and removed three proteins with over 50% missingness. The remaining 2,920 proteins were inverse rank normalized. We also obtained 30 standard biochemistry clinical markers (Data Field ID:17518) and haematological data included 31 full blood count indices (Data Field ID:100081).

### Multi-omic integration

We performed data integration using Multi-Omics Factor Analysis (MOFA) implemented in the *MOFA* R package^2^. Prior to model training, we applied feature selection to all four data modalities. For metabolomics, we selected 36 clinically validated metabolites previously shown to be predictive of disease outcomes^5^. For proteomics, we applied LASSO regression to each of the 2,920 proteins against 111 incident disease outcomes (both cancer and non-cancer) with ≥100 post-baseline cases, as defined by phecodes^6^. We only included diseases with ≥100 incident cases post-baseline. From each disease-specific model, we selected the three proteins with the largest absolute effect sizes, yielding a panel of 217 unique proteins.

For biochemistry, we retained 19 measurements after excluding 10 features already represented in the Nightingale metabolite panel (Albumin, Apolipoprotein A, Apolipoprotein B, Cholesterol, Creatinine, HDL cholesterol, LDL direct, Lipoprotein A, Triglycerides, Glucose) to avoid redundancy, and sex hormone-binding globulin, which was measured in both the biochemistry and proteomics assays. This resulted in a final set of 19 unique biochemistry markers. For haematology, we excluded percentage-based features due to collinearity with absolute counts, retaining 22 features.

We trained the MOFA model using the resulting 294 features across all modalities. All features were standardized (mean-centered and scaled to unit variance). We used a Gaussian likelihood for each data modality and applied spike-slab sparsity prior in the weights. The number of estimated latent factors was set to 20 to balance variance explained and model interpretability.

### Factor association with disease

To evaluate the relationship between latent factor and disease risk, we performed logistic regression with disease status as the binary outcome and individual factor scores as predictors. We included age and sex as covariates to adjust for demographic effects. We considered an association to be significant if it had a Bonferroni-corrected P-value of less than 0.05. That is,

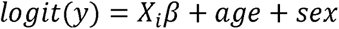

Where *y* ∈ (0,1) is the disease status and *X_i_*, i = 1,…, 20, represents the loading of the MOFA-derived factor for each individual. This is implemented in the *glm* function in R(v. 4.1.2).

### Factor association with polygenic risk scores

To investigate whether latent factors captured genetic liability for common traits, we analyzed associations between factor loadings and polygenic risk scores (PRS) across 36 traits, including 28 diseases and 8 quantitative traits. We used PRS values computed by Genomics PLC, derived from meta-analyses of external, non-UKB GWAS datasets^26^. We excluded their enhanced PRS that included UKB data to avoid circularity in analysis stemming from shared participants between the PRS derivation and target datasets. For each of the 36 traits, we obtained each individual’s PRS overlapping with the individuals in the UKB-PPP data (UKB Data Field ID: 301). We then performed linear regression of PRS and factor values in each disease including age and sex as covariates.

### Genome-wide association studies

For the factor-GWAS analysis, we included only individuals with European ancestry within the UKB-PPP cohort (N = 28,693, Data Field ID = 22006). We included genotyped SNPs that had minor allele frequency (MAF) > 0.01, Hardy Weinberg equilibrium (HWE) P-value > 1 x 10^-8^ and missingness rate < 10%, resulting 784,256 autosomal genotyped SNPs for the association testing.

For factor-GWAS, we fitted an additive linear regression model with each latent factor or disease as the outcome variable, including age, sex and the first 10 genetic principal components included as covariates using PLINK v2.0^31^.

To evaluate overlap with known trait-associated variants, we looked for overlap between factor-GWAS and signals reported in the NHGRI GWAS catalog^21^. We first extracted all SNPs in high linkage disequilibrium (LD, r^2^ > 0.8) with lead factor-GWAS SNPs reaching genome-wide significance (P < 5×10^-8^/20) using data from the 1000 Genomes Project (EUR panel). We then annotated these SNPs to genes using a 35 kb upstream and 10 kb downstream window and checked for overlap with GWAS catalog annotated genes.

### Gene set enrichment analysis

To identify biological pathways enriched in the genetic architecture of each latent factor, we performed gene set enrichment analysis using summary statistics from the factor-wide genome-wide association studies (factor-GWAS). We employed MAGMA^19^ to map SNPs to genes and compute gene-level association statistics for each of the 20 MOFA-derived latent factors. MAGMA accounts for linkage disequilibrium structure and gene-gene correlations when estimating gene-level effects. We then tested for enrichment of these gene-level associations across curated gene sets from the REACTOME pathway database^20^.

### Disease Prediction

To evaluate disease prediction performance, we selected the top 50 highest-weighted features from each latent factor and fitted logistic regression models with disease status as the outcome and age and sex as covariates in the 75% training cohort. Model discrimination was then evaluated in the 25% held-out test cohort. To obtain stable performance estimates, we performed 100 bootstrap resamples of the test set (5,000 individuals per resample) and computed the concordance index (C-index) for each iteration using the *Hmisc* R package^32^(version 5.2), which implements Harrell’s C-index as a measure of rank correlation between predicted and observed outcomes.

We only performed this analysis if there was a significant association between the factor and disease in the factor-disease association analysis. In comparison, we fitted the same regression model with all features in each single modality separately. We also fitted this logistic regression model for each feature separately, including features that were not used in the MOFA model (n=3,230). The rationale for this is that we wanted to investigate whether our method of feature selection performed better than single feature or single modality. To compare the different methods, we considered that they were statistically significant if the 95% confidence intervals did not intersect with each other.

### Validation with Test Set

To validate our results, we projected the weights of the model built on the 75% training set onto the 25% test set, computing factors for these individuals.

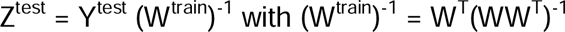

We then performed disease association analysis using the same covariates as for the training set. We also performed GWAS in the test set using these projected factors.

## Data availability

UKB data are available through a procedure described at https://www.ukbiobank.ac.uk/enable-your-research. Summary statistics from all analysis stages are included in Supplementary Files.

## Code availability

All code needed to reproduce all analyses, figures and tables are publicly available via GitHub https://github.com/yang-luo-lab/omicUKB

## Acknowledgements

We would like to gratefully acknowledge the UKB participants for their dedication to participating in ongoing research. This work and the incredible work of other UKB researchers would not have been possible without their kindness and dedication to the pursuit of medical science. UKB resource under approved application numbers 31295 and 30418.

## Author Contributions

Conceptualization, Formal Analysis, Software: E.S.N. and Y.L., R.N.

Project administration, Resources, Methodology: E.S.N, Y.L., J.T., L.J., J.B.

Writing original draft – E.S.N. and Y.L.

Writing-review and editing: E.S.N, Y.L., J.T., L.J., J.B.

---

**Supplementary Fig. 1.**
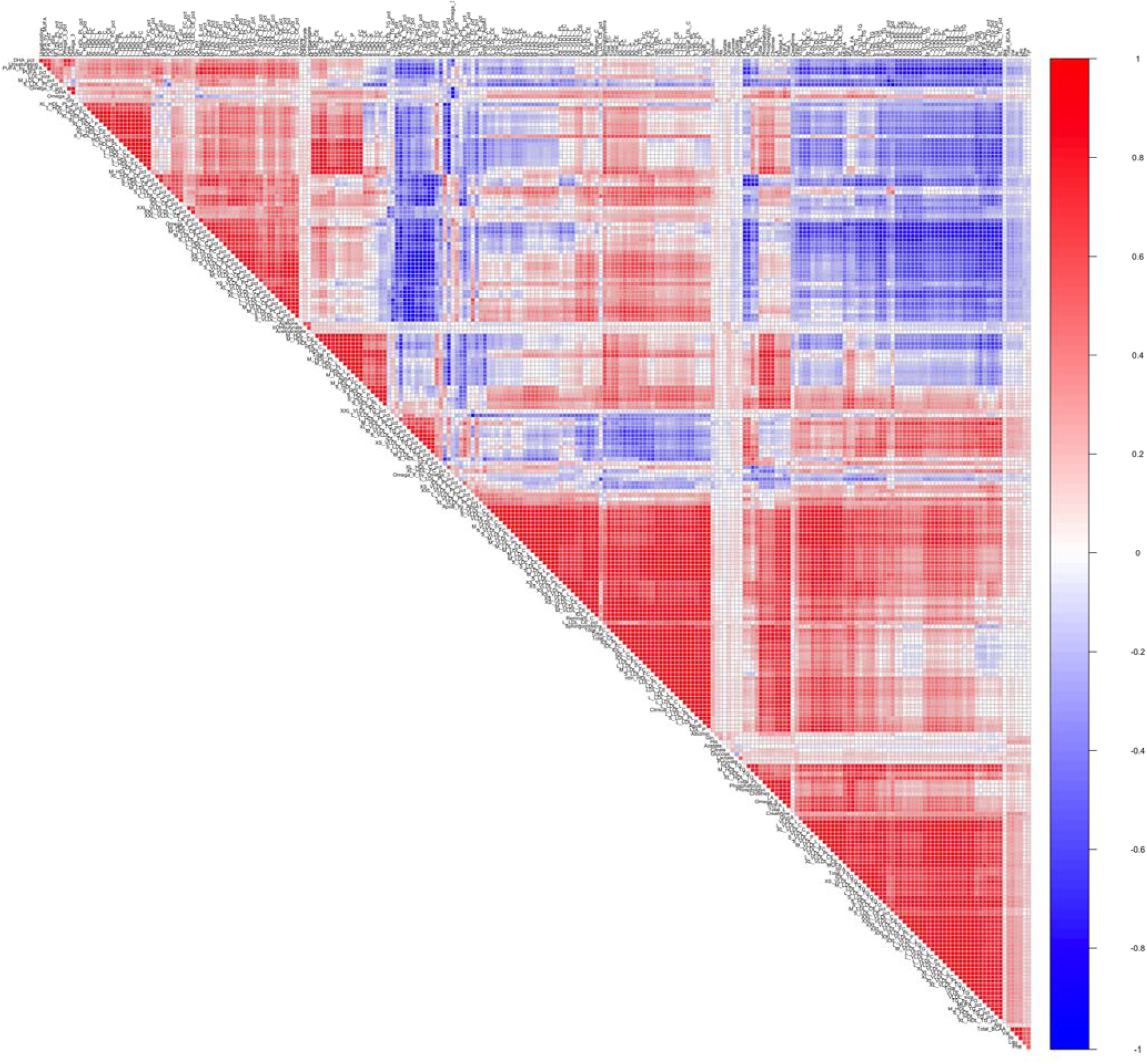
Correlation heatmap of metabolite levels in UKB. Pairwise Pearson correlation coefficients between metabolites are visualized as a color-coded heatmap, generated using the *R* package *corrplot* (v0.92). Metabolites are ordered by hierarchical clustering to highlight groups with similar correlation patterns. Blue and red indicate negative (r <0) and positive (r>0) correlations, respectively, with color intensity reflecting correlation strength. Only the upper triangle of the matrix is shown for clarity.

**Supplementary Fig. 2.**
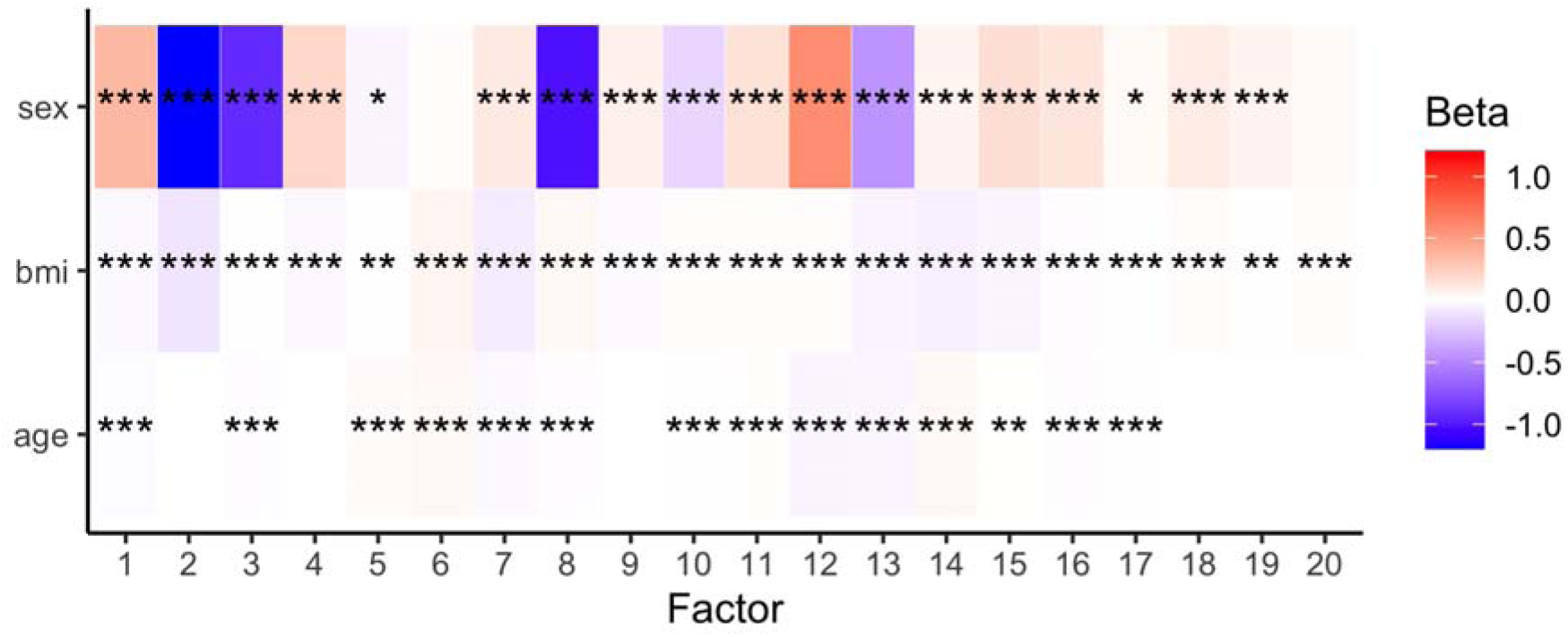
Associations between latent factors and demographic and anthropometric variables. Heatmap showing the beta coefficients from linear models associating each latent factor with age, sex, and BMI respectively. Colors indicate the direction and magnitude of association (blue = negative, red = positive, white = no association). Significance is denoted by asterisks (*CPC<C0.05/20/3, **CPC<C0.01/20/3, ***CPC<C0.001/20/3; Bonferroni-corrected).

**Supplementary Fig. 3.**
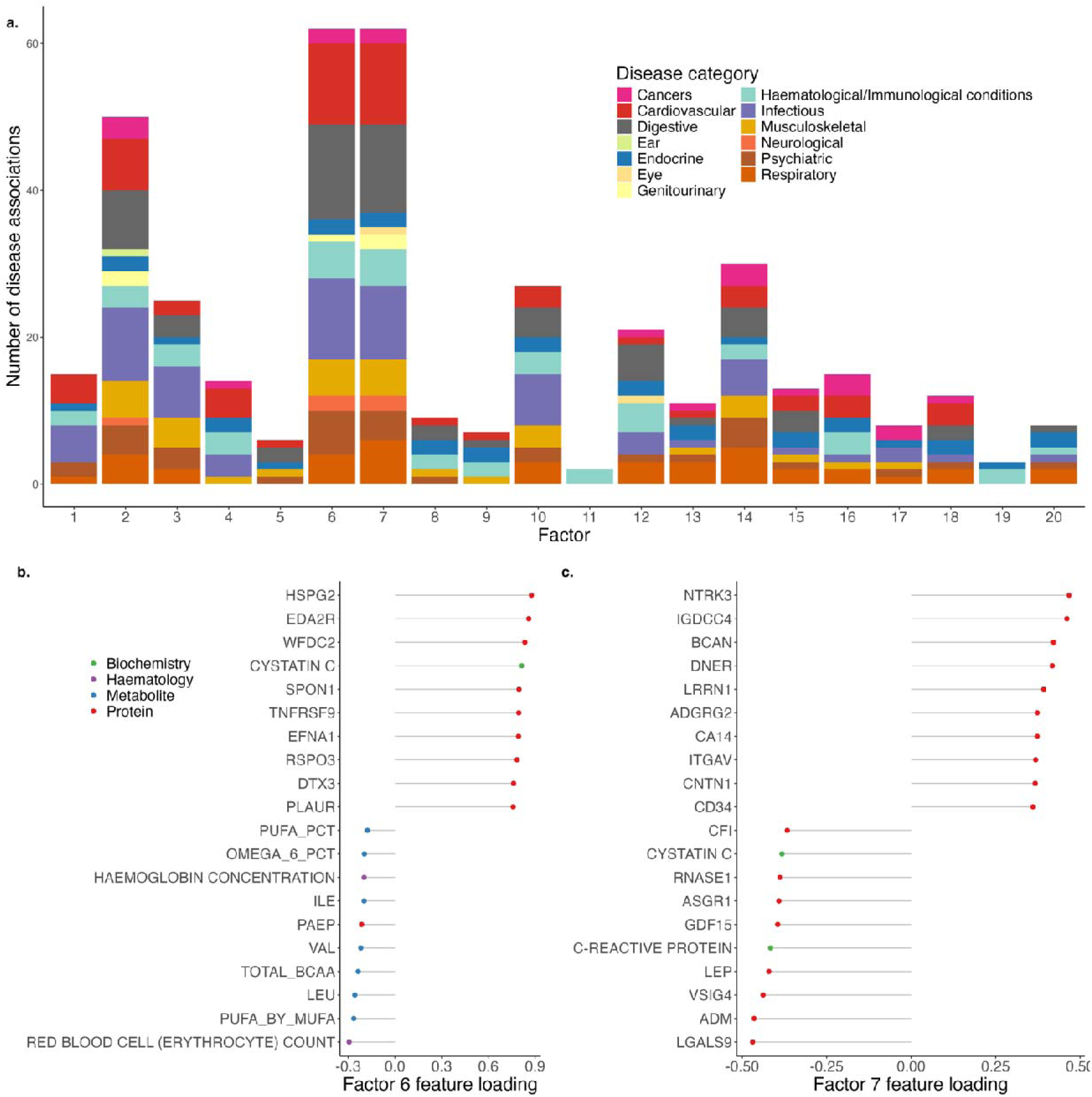
Number of significant disease associations for each factor. **a.** Number of significant disease associations per factor (Bonferroni-corrected p < 0.05). Bars are colored by disease category, showing that Factors 6 and 7 have the highest number of disease associations across multiple categories. Top 10 positive and negative feature loadings for (**b**) Factor 6 and (**c**) Factor 7, colored by omics modality (metabolites in blue, biochemistry in green, proteins in red, haematology in purple). Factor 6 shows strong loadings across multiple omics types. Factor 7 exhibits distinct feature patterns compared to Factor 6, with a strong proteomic contribution.

**Supplementary Fig. 4.**
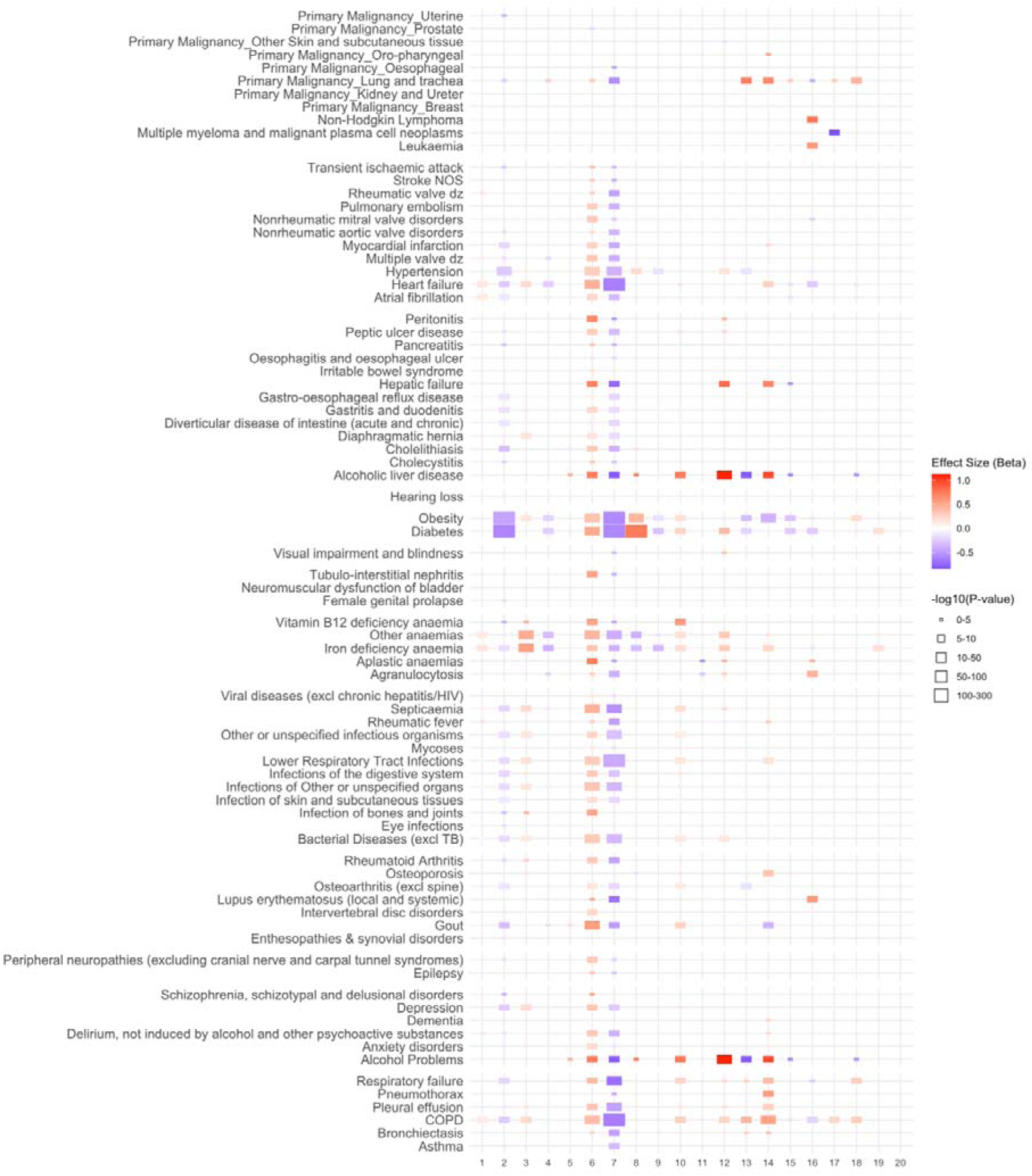
Factor-disease associations for 111 incident diseases in the UKB-PPP. The effect sizes and P-values are estimated via logistic regression adjusted for age and sex. Tiles are colored based on association strength (regression), with size proportional to -log_10_(P-value).

**Supplementary Fig. 5.**
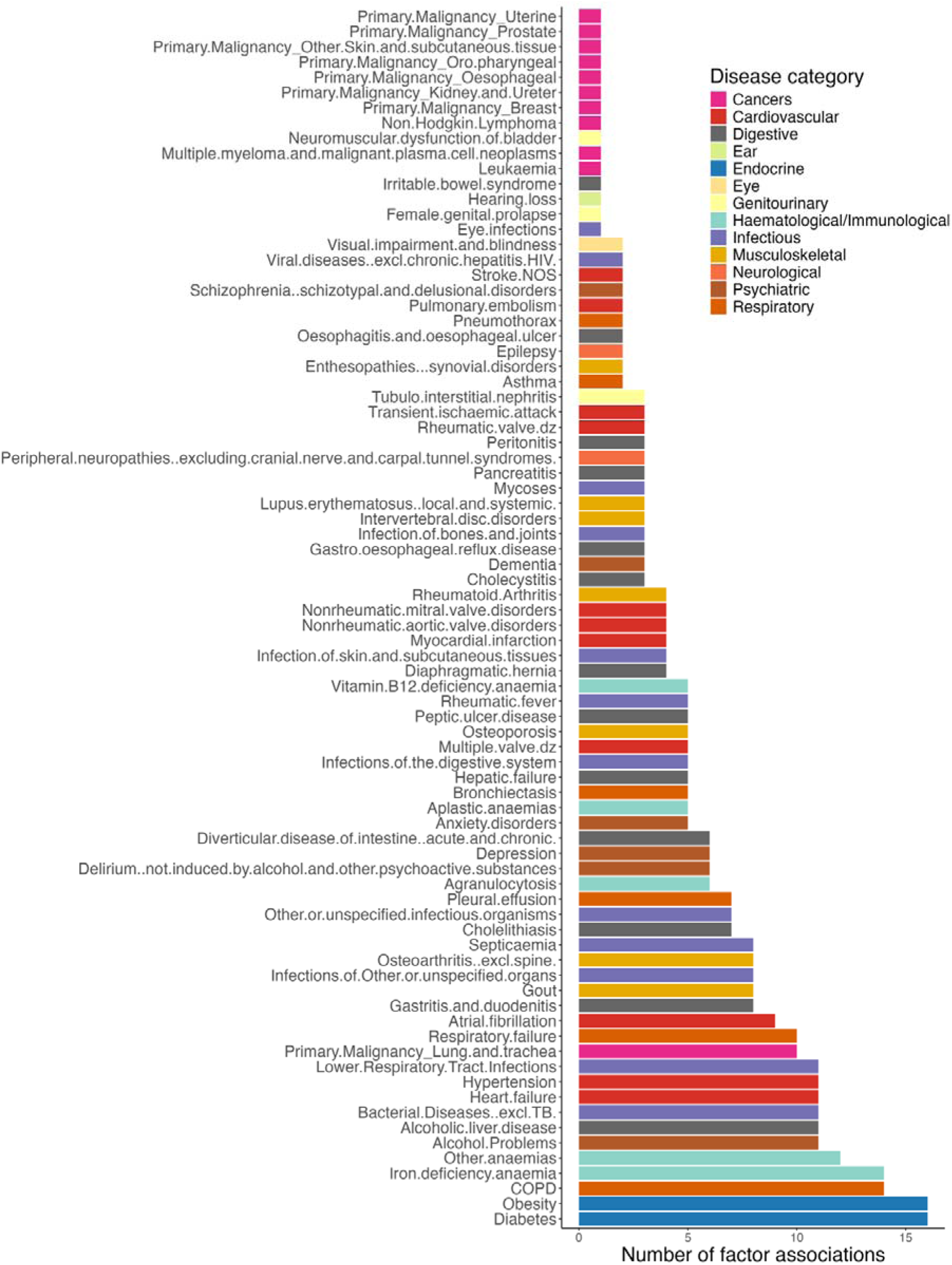
Number of significant factor associations for each disease category. Bar plot showing the number of latent factors significantly associated with each disease, Colored by disease category. Diseases are ordered by the total count of associated factors.

**Supplementary Fig. 6.**
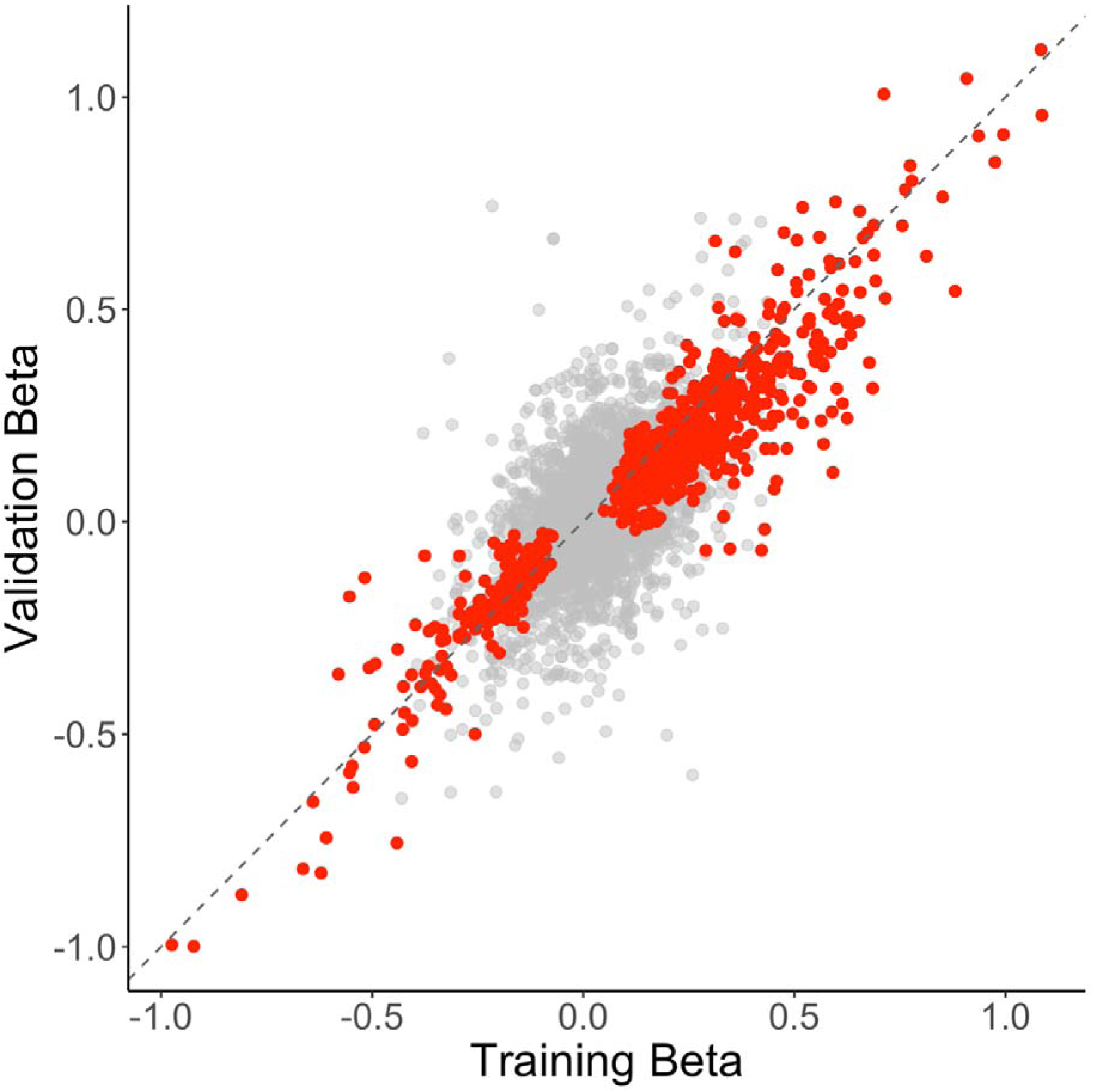
Scatter plot of training versus validation effect sizes (betas) for factor-disease associations. Each point represents a factor-disease pair. Grey points indicate associations not reaching Bonferroni significance in the discovery set (discovery P ≥ 0.05/111/20), while red points indicate significant associations (discovery P < 0.05/111/20). The dashed line represents the identity line (y = x). We observed a high degree of replication, with 98.5% (394 of 400) of the discovery associations showing a concordant direction of effect in the validation cohort.

**Supplementary Fig. 7.**
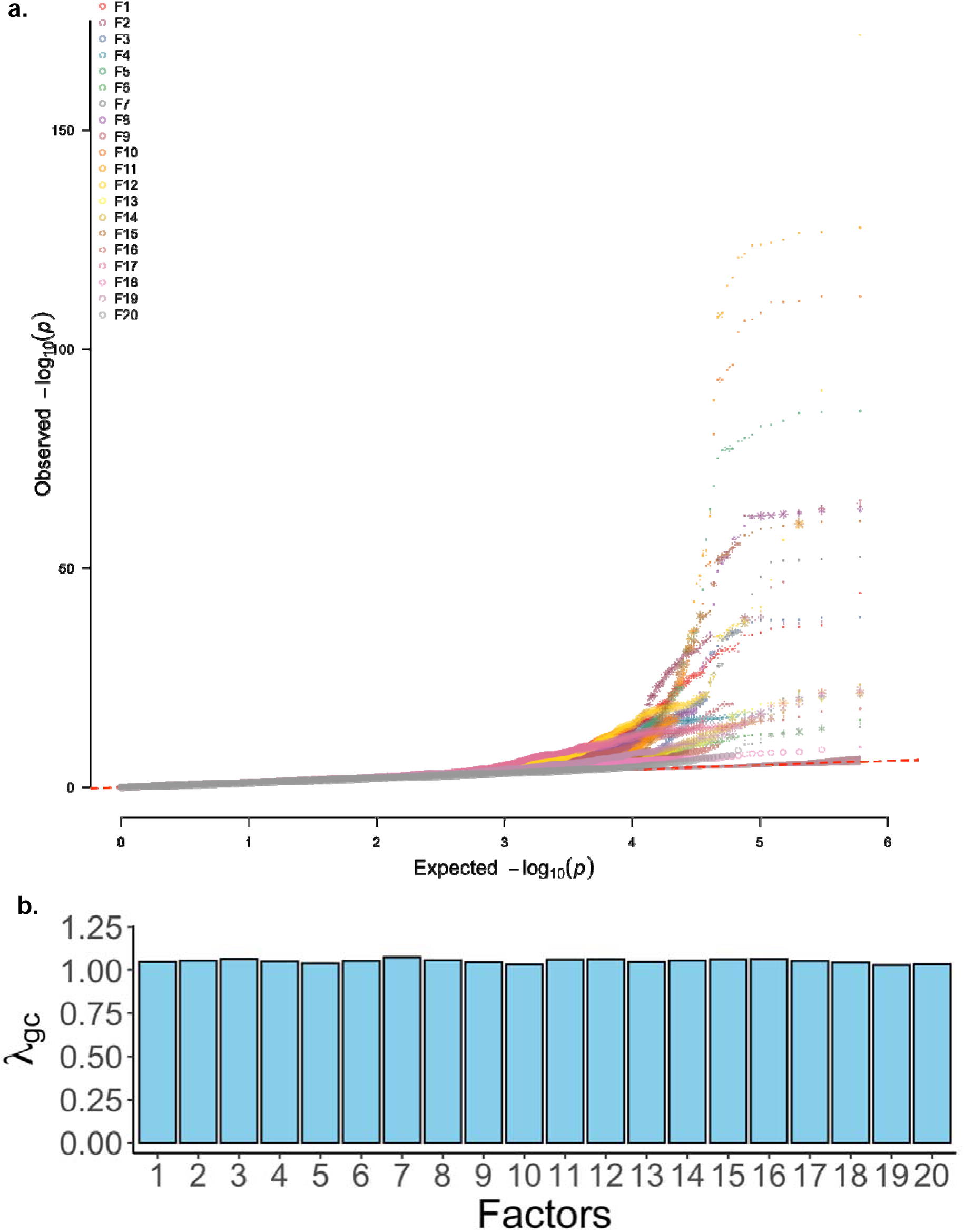
Genomic Inflation in factor-GWAS analyses. **a.** quantile-quantile (QQ) plots for GWAS of each factor. Each plot compares the observed versus expected distribution of p-values under the null hypothesis for all tested variants. **b.** Barplot shows the λ_gc_ values for all 20 factors, indicating the degree of test statistic inflation in each genome-wide association analysis.

**Supplementary Fig. 8.**
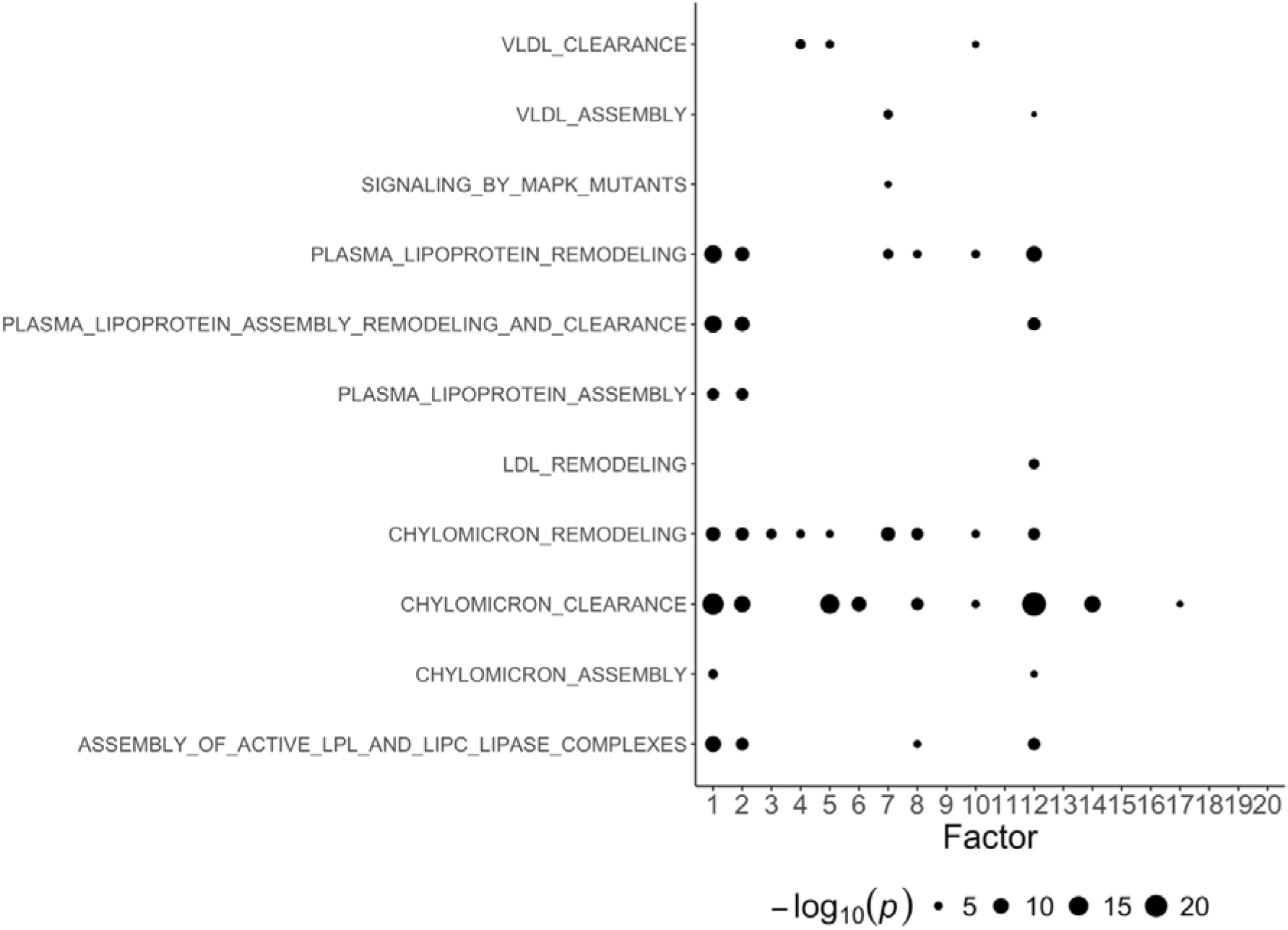
Reactome pathway analysis of factor GWAS using MAGMA gene-set enrichment. Each point represents a Reactome pathway, with size indicating the statistical significance (−log_10_(P)) of enrichment for genes associated with each factor.

**Supplementary Fig. 9.**
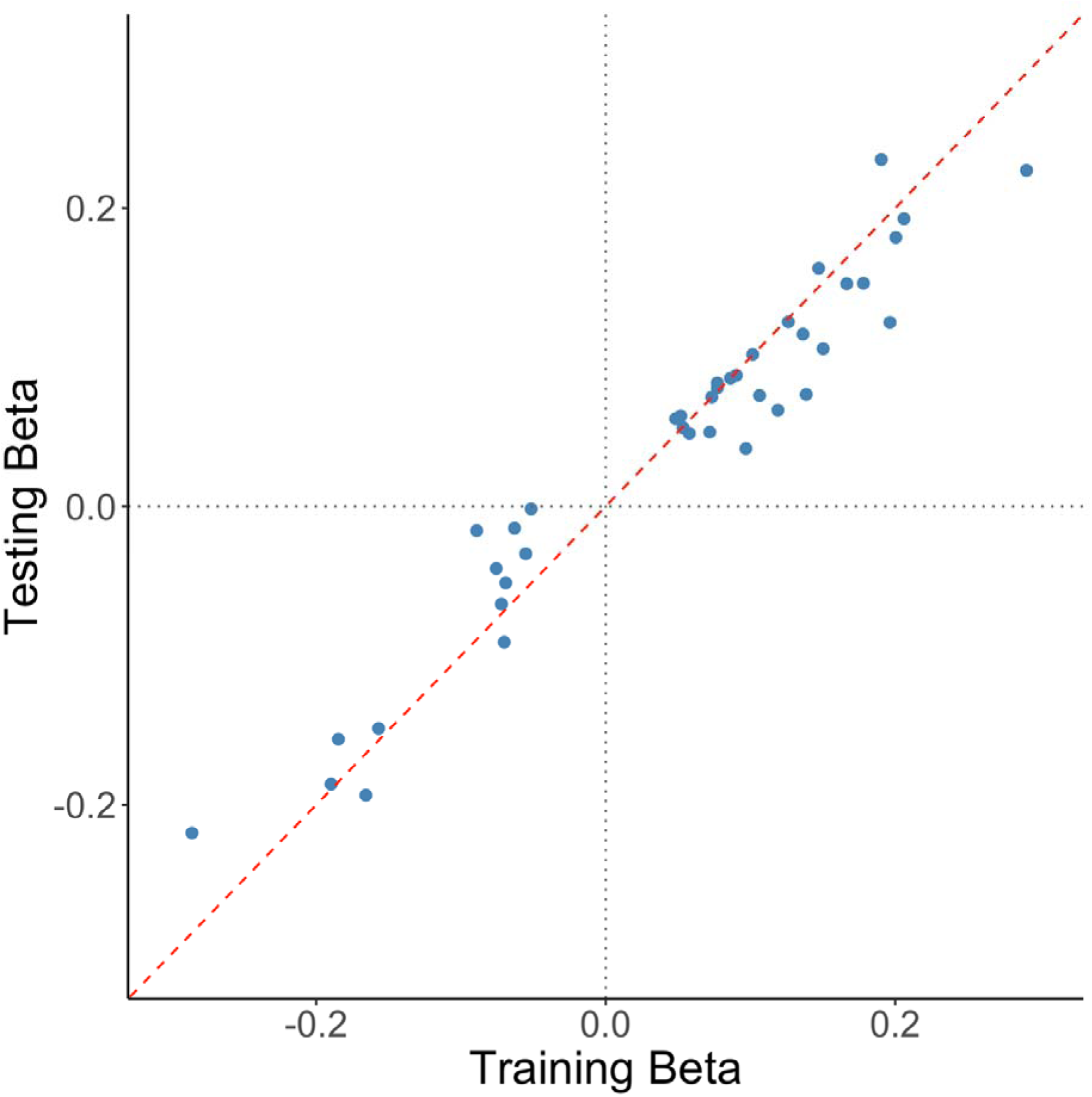
Beta values from the training set compared to beta values from the 25% held-out test set. Each point represents an independent variant from the factor-GWAS. The red dashed line indicates the identity line (y = x), and gray dotted lines mark the zero reference for both axes. Out of the 37 lead SNPs in the 75% training set, all of them showed the same direction of effect in the test set, all of which had P-values < 0.05.

**Supplementary Fig. 10.**
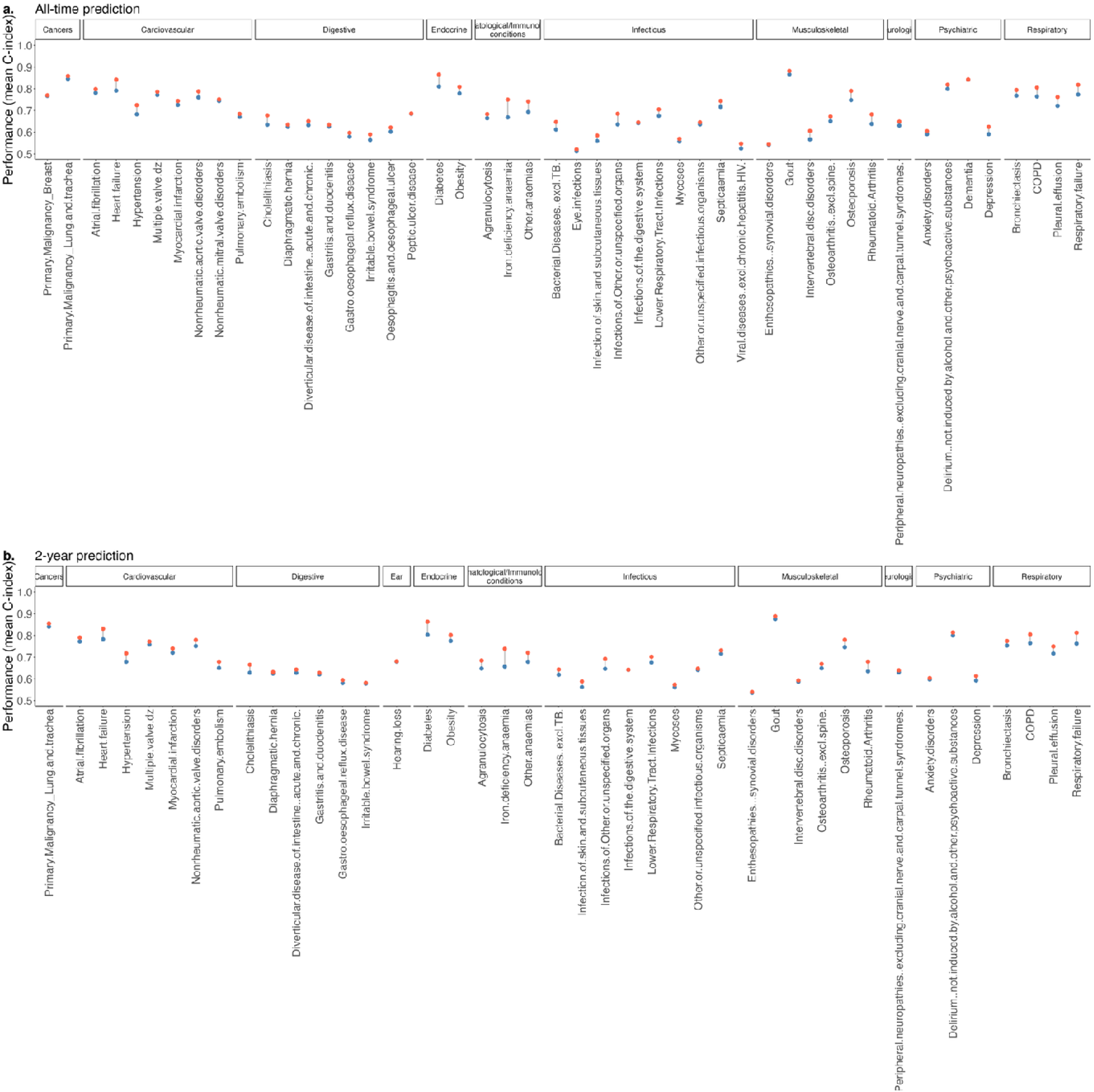
Imroved predictive performance of common diseases using multi-omic models compared to single-biomarker models. Increase in C-index when using models trained on 50-omic features (orange) versus a single biomarker (blue) across **a**. 48 common incident diseases and **b.** 42 common incident disease two-year after blood draw.

